# Determinants of Prehospital Management of Superficial Burns in Pediatric Patients at Health Facilities in Mazabuka District: A Cross-Sectional Study

**DOI:** 10.64898/2026.11.05.26367398

**Authors:** Davies Kasoma, Heather E. Rosen, Penelope Kantu Machona, Choolwe Jacobs, Patrick Musonda

## Abstract

**Background:** Burn injuries in children are common in low-resource settings, yet little is known about how they are managed before reaching health facilities in Zambia. Early care at home plays a key role in recovery, but harmful practices are still widely used. We examined how caregivers manage superficial burns in children and identified factors linked to good prehospital care in Mazabuka District.

**Methods:** We conducted a cross-sectional study between September and December 2025 among 337 caregivers of children aged 0–15 years presenting with superficial burns. The study was conducted at three health facilities in Mazabuka District. Data were collected using structured interviews and clinical records. Prehospital burn care was classified as good or poor based on the recommended first-aid practices. Associations were assessed using Firth penalised logistic regression, and results are reported as adjusted odds ratios (AORs) with 95% confidence intervals (CIs).

**Results:** Only 13.1% (44/337) of caregivers provided good prehospital burn care. Urban residence was associated with lower odds of good management (AOR 0.22; 95% CI: 0.05–0.92; p=0.038). Older caregiver age reduced the odds of good care (AOR 0.92; 95% CI: 0.86–0.98; p=0.011), while increasing child age improved it (AOR 1.25; 95% CI: 1.10–1.43; p<0.001). Longer time to reach a facility was also associated with poorer care (AOR 0.96; 95% CI 0.94–0.98; p<0.001).

**Conclusion:** Good prehospital burn care was uncommon. It was associated with time to the health facility, caregiver age, child age, and urban versus rural residence, rather than with proximity to a facility alone. Programmes should promote cooling with running water, avoidance of harmful home applications, and prompt care-seeking. Messages in routine child health services may reach caregivers of young children; additional approaches are needed for older caregivers and urban communities.

## Introduction

Pediatric burn injuries are a serious global health problem and a major cause of mortality and morbidity among children[1, 2]. In 2019, 7.09 million pediatric burn cases occurred globally, including 1.66 million among children under five years.[3, 4]. These injuries constitute an important public-health burden in the WHO African Region, a low- and middle-income country (LMIC) context in which their epidemiology, sex distribution, and emergency-department outcomes have been synthesised in a single systematic review[5].

Evidence on burns care in Zambia remains limited and is concentrated in a few healthcare facilities. Available studies primarily describe patient characteristics, burn mechanisms, complications and clinical outcomes, while gaps in burns research and community burn-prevention knowledge remain[6, 7]. Effective prehospital care is critical for reducing burn-related complications and improving outcomes[8]. Effective initial management includes cooling the burn with running water for at least 10–20 minutes, covering the wound with a clean dressing, and transporting the child promptly to a health facility[8, 9]. These steps help reduce tissue damage, limit the risk of infection, and reduce the need for surgery.

Despite the widespread nature of the problem, appropriate prehospital burn care is rarely practiced in sub-Saharan Africa. Traditional burn care, including plant and animal products, remains widely used in sub-Saharan Africa[10]. Caregivers more commonly applied harmful substances including oils, herbs, or animal products to the wound[11]. The primary driver of poor care is believed to be a lack of knowledge among first responders, who are most often family members rather than trained health workers[11]. There is a lack of evidence on the adequacy of prehospital care for children with burns in Zambia, and factors associated with appropriate prehospital care have not been previously studied. Therefore, the objective of this study was to determine how caregivers manage superficial burns in children and factors linked to good prehospital care in Mazabuka District. The study was designed to inform targeted interventions for early pediatric burn care in Zambia and similar settings.

## Methods

### Study Design and Setting

This cross-sectional analytical study was conducted at three health facilities in Mazabuka District, Southern Province, Zambia: Mazabuka General Hospital (the principal district referral facility), Kaonga Mini Hospital, and Nakambala Urban Clinic. Mazabuka District lies approximately 127 km south of Lusaka and has a population of slightly over 250,000 (ZamStats, 2022 Census). The facilities collectively serve urban and peri-urban populations and represent the main points of first clinical contact for pediatric burn injuries in the district. Data were collected between 25 September and 14 December 2025. The study is reported in line with STROBE[12].

### Participants, Eligibility, and Sampling

The sample comprised primary caregivers and their children aged 0–15 years presenting with superficial burns within seven days of injury. Caregivers of children with deep partial or full-thickness burns, those presenting without a caregiver, and those who declined consent were excluded. The minimum sample size was calculated using Cochran’s formula (Z = 1.96, P = 0.44, d = 0.05), yielding n = 379. Participants were recruited by systematic sampling: based on an anticipated number of eligible episodes, every third eligible caregiver presenting at any of the three facilities was invited. Respondents were primary caregivers of children aged 0–15 years, with the children serving as index patients. Using Cochran’s formula, the minimum required sample size was 379. The figure of 360 represented the anticipated number of eligible presentations and was used only to determine a 1-in-3 sampling interval; it was not an alternative sample size. Recruitment continued throughout the study period, during which 337 eligible caregivers were enrolled

### Data Collection and Instruments

A structured, interviewer-administered questionnaire was developed by adapting items from validated prehospital burn-care guidelines WHO[13] and pre-tested on approximately 20 participants from the final study sample. The questionnaire captured caregiver sociodemographic characteristics, burn-event details, prehospital care practices, health-seeking behaviour, transport and distance to facility, and caregiver beliefs regarding traditional remedies. In addition, a standardised clinical record abstraction form was used to extract burn characteristics and clinical status at arrival from patient medical records. Following stabilisation of the injured child, eligible caregivers were approached by trained research assistants and interviewed in the local language using password-protected tablet devices. Daily completeness and consistency checks were conducted throughout the data-collection period. The pre-test involved approximately 20 caregivers at the same three Mazabuka facilities to check language, flow, and timing; those interviews were excluded from the analysis, and items were revised.

### Study Variables

The primary outcome was prehospital burn wound management, classified as good or poor. Poor management was defined as failure to cool the burn with running water for at least 20 minutes cover the wound with a clean dressing and/or apply harmful substances (e.g., butter, oil, toothpaste, ash, or herbal preparations) before facility presentation. Explanatory variables were organised across four domains of Andersen’s Behavioural Model of Health Services Use[14] predisposing factors (caregiver age, education, child age, and sex); enabling factors (transport mode, distance to facility, first responder presence at a time of incident, and time to facility); need factors (burn type); and health behavioural factors(first aid practices for the immediate management of superficial burns such as cooling the burn with running water and avoiding harmful applications and caregiver beliefs in traditional remedies). Belief in traditional remedies was measured on a five-point Likert scale (strongly disagree to strongly agree) and collapsed as agree/strongly agree versus neutral/disagree/strongly disagree.

### Data Management

Data were collected on password-protected devices and transferred daily to a secure server accessible only to authorised personnel. Participants were anonymised using unique identification codes. Before analysis, data were checked for completeness and logical consistency; discrepancies were resolved by the field supervisor. Double-entry verification was performed on a random 10% subsample to detect transcription errors.

### Statistical Analysis

All analyses were performed in Stata version 17 (StataCorp, TX, USA). Categorical variables were summarised as frequencies and proportions; continuous variables as medians and interquartile ranges (IQR). Group comparisons used chi-square or Fisher’s exact tests for categorical variables and the Wilcoxon rank-sum test for continuous variables. Unadjusted associations were estimated by standard logistic regression, producing crude odds ratios (ORs) with 95% confidence intervals. For the adjusted model, Firth penalised likelihood logistic regression was used to address sparse-data and near-separation problems inherent in the data [15]. Two variables, first aid administered and any first responder, were excluded from the multivariable model due to perfect or near-perfect separation; traditional remedy belief was excluded due to quasi-complete separation. Traditional remedy belief was excluded from the multivariable model because of quasi-complete separation and is presented only as a descriptive comparison in Table 1; a standalone regression was not fitted.

**Table 1.** Characteristics of caregivers and children presenting with superficial burns, Mazabuka District1.

| Characteristic | Total (n = 337) | Poor management<br>(n = 293) | Good management<br>(n = 44) | p value |
|---|---|---|---|---|
| **Caregiver sex** |  |  |  | 0.349* |
| Female | 291 (86.4%) | 255 (87.0%) | 36 (81.8%) |  |
| Male | 46 (13.6%) | 38 (13.0%) | 8 (18.2%) |  |
| **Sex of child** |  |  |  | 0.395* |
| Female | 116 (34.4%) | 98 (33.4%) | 18 (40.9%) |  |
| Male | 221 (65.6%) | 195 (66.6%) | 26 (59.1%) |  |
| **Residence** |  |  |  | 0.650* |
| Rural | 49 (14.5%) | 44 (15.0%) | 5 (11.4%) |  |
| Urban | 288 (85.5%) | 249 (85.0%) | 39 (88.6%) |  |
| **Caregiver education** |  |  |  | 0.047* |
| No formal education | 30 (8.9%) | 26 (8.9%) | 4 (9.1%) |  |
| Primary | 95 (28.2%) | 89 (30.4%) | 6 (13.6%) |  |
| Secondary | 173 (51.3%) | 148 (50.5%) | 25 (56.8%) |  |
| Tertiary | 39 (11.6%) | 30 (10.2%) | 9 (20.5%) |  |
| **Burn type** |  |  |  | 0.903* |
| Contact | 131 (38.9%) | 114 (38.9%) | 17 (38.6%) |  |
| Flame | 23 (6.8%) | 21 (7.2%) | 2 (4.5%) |  |
| Scald | 183 (54.3%) | 158 (53.9%) | 25 (56.8%) |  |
| **Transport to facility** |  |  |  | 0.307* |
| Walk or bicycle | 149 (44.2%) | 130 (44.4%) | 19 (43.2%) |  |
| Private motorised | 21 (6.2%) | 16 (5.5%) | 5 (11.4%) |  |
| Public transport | 167 (49.6%) | 147 (50.2%) | 20 (45.5%) |  |
| **First aid administered‡** |  |  |  | 0.002* |
| No | 45 (13.4%) | 45 (15.4%) | 0 (0.0%) |  |
| Yes | 292 (86.6%) | 248 (84.6%) | 44 (100.0%) |  |
| **Any first responder‡** |  |  |  | 0.143* |
| No | 17 (5.0%) | 17 (5.8%) | 0 (0.0%) |  |
| Yes | 320 (95.0%) | 276 (94.2%) | 44 (100.0%) |  |
| **Traditional remedy belief§** |  |  |  | <0.001* |
| Agree / strongly agree | 249 (73.9%) | 247 (84.3%) | 2 (4.5%) |  |
| Neutral / disagree / strongly disagree | 88 (26.1%) | 46 (15.7%) | 42 (95.5%) |  |
| **Distance to facility** |  |  |  | 0.235* |
| Short (≤5 km) | 292 (86.6%) | 251 (85.7%) | 41 (93.2%) |  |
| Long (>5 km) | 45 (13.4%) | 42 (14.3%) | 3 (6.8%) |  |
| **Caregiver age, years, median (IQR)** | 32.0 (28.0 to 39.0) | 32.0 (28.0 to 40.0) | 32.5 (29.0 to 36.0) | 0.353** |
| **Child age, years, median (IQR)** | 5.0 (3.0 to 7.0) | 5.0 (3.0 to 7.0) | 6.5 (4.0 to 9.0) | 0.056† |
| **Time to facility, minutes, median (IQR)** | 25.0 (15.0 to 45.0) | 25.0 (15.0 to 50.0) | 15.0 (10.0 to 20.0) | <0.001** |

Data are n (%) or median (IQR). p values compare poor versus good management. \*χ² or Fisher’s exact test for categorical variables (Fisher’s exact test when any expected cell count was <5). \*\*Wilcoxon rank-sum test for continuous variables.

### Ethical Considerations

Ethical approval was obtained from the University of Zambia Biomedical Research Ethics Committee (UNZABREC REF. No. 6845-2025), with institutional permission from the National Health Research Authority (NHRA-2425/07/07/2025), and site-level authorisation from Mazabuka District Health Management. Written informed consent was obtained from all caregivers prior to data collection. Participants were assured of their right to withdraw without consequence and of the confidentiality of their responses. All data were anonymised and stored securely. The study was conducted in accordance with the Declaration of Helsinki[16].

## Results

### Participant’s characteristics

A total of 337 caregivers were included, of whom 13.1% (n=44/337) had good prehospital management. Most caregivers were female (86.4%, n=291/337), and most children were male (65.6%, n=221/337). Most families lived in urban areas (85.5%, n=288/337) and were within 5 km of the facility (86.6%, n=292/337). Scald burns were the most common injury type (54.3%, n=183/337), while public transport was the most common means of reaching the facility (49.6%, n=167/337). Most caregivers administered first aid (86.6%, n=292/337), including all caregivers in the good-management group (100%, n=44/44) compared with 84.6% (n=248/293) in the poor-management group. Education level differed between groups (*p* = 0.047). Traditional remedy beliefs also differed: 84.3% (n=247/293) of caregivers in the poor-management group agreed or strongly agreed that traditional remedies are appropriate, compared with 4.5% (n=2/44) in the good-management group *(p* < 0.001). Median time to the facility was 15 minutes (IQR 10–20) in the good-management group and 25 minutes (IQR 15–50) in the poor-management group (p < 0.001). Caregiver sex, child sex, residence, burn type, transport, first responder, distance to the facility, caregiver age and child age were not significantly associated with management outcome (*p* > 0.05).

### Factors associated with burn management

After adjusting for all other variables in the model, four factors were independently and significantly associated with good burn wound management (Table 2). Urban residence was associated with lower odds of good management compared to rural residence (AOR 0.22; 95% CI: 0.05–0.92). Each additional year of caregiver age was associated with slightly lower odds (AOR 0.92; 95% CI: 0.86–0.98). Conversely, each additional year of child age was associated with higher odds of good management (AOR 1.25; 95% CI: 1.10–1.43). Finally, each additional minute taken to reach the facility was associated with lower odds of good management (AOR 0.96; 95% CI: 0.94–0.98). No other variable reached statistical significance in the adjusted model.

**Table 2.** Factors associated with good prehospital burn wound management2.

| **Characteristic** | **Crude OR (95% CI)** | **p value** | **Adjusted OR\* (95% CI)** | **p value** |
|---|---|---|---|---|
| **Caregiver sex** |  |  |  |  |
| Female (reference) | 1.0 |  | 1.0 |  |
| Male | 1.49 (0.64 to 3.45) | 0.350 | 1.75 (0.72 to 4.29) | 0.219 |
| **Sex of child** |  |  |  |  |
| Female (reference) | 1.0 |  | 1.0 |  |
| Male | 0.73 (0.38 to 1.39) | 0.333 | 0.77 (0.39 to 1.52) | 0.445 |
| **Residence** |  |  |  |  |
| Rural (reference) | 1.0 |  | 1.0 |  |
| Urban | 1.38 (0.51 to 3.69) | 0.523 | 0.22 (0.05 to 0.92) | 0.038 |
| **Caregiver education** |  |  |  |  |
| No formal education (reference) | 1.0 |  | 1.0 |  |
| Primary | 0.44 (0.11 to 1.67) | 0.227 | 0.28 (0.07 to 1.15) | 0.077 |
| Secondary | 1.10 (0.35 to 3.42) | 0.872 | 0.67 (0.19 to 2.39) | 0.540 |
| Tertiary | 1.95 (0.54 to 7.08) | 0.310 | 0.86 (0.20 to 3.68) | 0.841 |
| **Burn type** |  |  |  |  |
| Contact (reference) | 1.0 |  | 1.0 |  |
| Flame | 0.64 (0.14 to 2.97) | 0.568 | 0.79 (0.17 to 3.61) | 0.764 |
| Scald | 1.06 (0.55 to 2.06) | 0.861 | 0.98 (0.47 to 2.01) | 0.948 |
| **Transport to facility** |  |  |  |  |
| Walk or bicycle (reference) | 1.0 |  | 1.0 |  |
| Private motorised | 2.14 (0.70 to 6.51) | 0.181 | 1.48 (0.42 to 5.15) | 0.541 |
| Public transport | 0.93 (0.48 to 1.82) | 0.834 | 0.75 (0.35 to 1.58) | 0.445 |
| **Distance to facility** |  |  |  |  |
| Short (≤5 km) (reference) | 1.0 |  | 1.0 |  |
| Long (>5 km) | 0.44 (0.13 to 1.48) | 0.183 | 1.70 (0.40 to 7.15) | 0.471 |
| **Caregiver age, per 1 year** | 0.97 (0.93 to 1.01) | 0.131 | 0.92 (0.86 to 0.98) | 0.011 |
| **Child age, per 1 year** | 1.08 (0.98 to 1.19) | 0.104 | 1.25 (1.10 to 1.43) | <0.001 |
| **Time to facility, per 1 minute** | 0.97 (0.95 to 0.99) | 0.003 | 0.96 (0.94 to 0.98) | <0.001 |

\*Adjusted odds ratios from penalised logistic regression (n = 337). Crude odds ratios from standard maximum-likelihood logistic regression. CI = confidence interval; OR = odds ratio. First aid administered, any first responder, and traditional remedy belief were excluded from the multivariable model because of perfect or quasi-complete separation.

## Discussion

In our study of caregivers in Mazabuka District, good prehospital wound management was uncommon, with only 13% meeting our definition of good care. Urban residence, older caregiver age, and longer time to a health facility were associated with poorer management, while older child age was associated with better management. Overall, our findings point to important gaps in the early care of childhood burns before children reach a health facility. These findings highlight the need to strengthen simple, timely, and appropriate burn first-aid practices at community level

The low rate of good prehospital wound management in Mazabuka aligns with patterns observed across sub-Saharan Africa. In a study from South Africa, over 90% of children did not receive any prehospital wound care; only a few received water irrigation or appropriate dressings [17]. Research indicates that even when water is used to clean the wound, it is often for too short a time or combined with other substances like eggs or toothpaste that have no proven benefit [17–19].Similar trends are seen in Malawi and Nigeria, where traditional remedies such as eggs or pap are commonly used instead of water[18, 20]. These findings highlight a persistent gap between what is recommended and what actually happens at home or in the community before reaching medical care. Our study assessed first-aid practices using a WHO-based knowledge score. Although most caregivers reported providing first aid, only 13% met our definition of good care. This gap between reported practice and recommended care may reflect incomplete knowledge, competing beliefs, or both.

Caregivers with belief in traditional remedies were significantly more likely to be in the poor management group. Similar findings were reported in studies from Malawi and South Africa, where many caregivers either provided no pre-hospital burn care or relied on traditional remedies due to cultural beliefs, misconceptions, and limited knowledge of appropriate burn first aid [17, 18]. In Malawi and other African countries, eggs, oil, honey, toothpaste, and herbal mixtures are frequently applied to burns because they are believed to prevent blisters or promote healing [17, 18, 20]. We did not include traditional-remedy beliefs in the multivariable model. However, caregivers with poor management were more likely to view traditional remedies as appropriate than those with good management.

These beliefs are passed down through families and communities rather than taught by health workers [18]. The social legitimacy of traditional remedies makes it hard for health messages from clinics to change behavior quickly. Studies indicate that health programs that work with community leaders and respect local beliefs may be more successful than those that simply provide information about correct techniques[18, 21].

Shorter time from injury to arrival at a health facility is linked to better prehospital wound management [22, 23]. Delays can be due to distance but also because families try home remedies before seeking formal care [22, 23]. Even when families live close to a clinic or hospital, they may wait too long if they believe home treatment is effective. Education campaigns should stress the importance of seeking medical help immediately after a burn rather than trying multiple home treatments first [24]. In our study, longer time from injury to facility arrival was associated with lower odds of good prehospital management. This finding reflects prehospital care practices, not clinical outcomes, as healing, grafting, and death were not measured

Surprisingly, living in an urban area was not associated with better prehospital burn care in this study. Previous study also found that urban caregivers have lower odds of providing good first aid compared to rural caregivers after adjusting for other factors [25]. Similar gaps in caregiver knowledge and practice have been reported in an urban African burn-unit sample[26]. This may be because urban areas offer easier access to informal markets selling unproven remedies or because busy lifestyles reduce supervision and prompt action after injuries [25]. Urban residence alone may not ensure appropriate prehospital burn care, as caregiver knowledge, beliefs, and access to accurate burn first-aid information appear to be more important determinants of practice than place of residence. Therefore, burn first-aid education should target both urban and rural populations[27]. In our study, urban residence was associated with lower adjusted odds of good management than rural residence, although most participants were urban (85.5%).

Our study found that older caregivers were less likely to provide good prehospital wound management. This may be because older adults rely more on traditional practices learned earlier in life and may not have been reached by recent health campaigns using mobile phones or community outreach about burn care [22, 25] Older children may receive better prehospital burn care than infants and toddlers because caregivers often find it more difficult to manage burns in very young children. Young children may resist cooling with running water due to distress, and some caregivers may incorrectly believe that cooling or pain relief is unsafe for them. These challenges and knowledge gaps may result in less effective first aid before reaching the hospital [22].Caregivers of very young children may benefit from improved burn first-aid education, although further research is needed to determine its impact on prehospital care.

The modelled findings (time to facility, caregiver age, child age, urban residence) and the descriptive belief contrast suggest that caregiver actions after a burn, rather than lack of a nearby clinic, may shape early care[28]. Existing Community Health Assistant, outreach, antenatal, and under-five platforms can deliver cool-water, no-harmful-application, and early-attendance messages; older caregivers and urban communities may need additional channels important gaps remain.

### Strengths and Limitations

Our study adds useful, real-world evidence from a Zambian district where little is published on how burns are managed before reaching a health facility, helping to fill an important gap in the region. The study recruited patients from three facilities which strengthens the reliability of the findings. That said, the results should be interpreted with caution because our study was cross-sectional and cannot establish causality, and the reliance on self-reported practices may have introduced social desirability bias, whereby some caregivers may have overstated their adherence to recommended childcare practices[29]In addition, our study included only those who came to health facilities, likely missing those with the poorest care practices, and did not capture factors like income, health literacy, or the exact home treatments used, which limits how fully the results can explain what is happening in practice. Another limitation of this study was the relatively small sample size, which did not meet the calculated minimum and may have reduced the statistical power of the analysis. This was primarily due to seasonal variation in injury occurrence, with more cases presenting in the winter and substantially fewer in the summer, which limited participant recruitment during the study period. The small number of outcome events also resulted in complete or quasi-complete separation for some variables, limiting their inclusion in the multivariable model and potentially affecting the stability and precision of the estimated associations[30]. The calculated minimum sample size was 379; 337 caregivers were enrolled because eligible volume in the study window was lower than the 360 presentations assumed for the sampling interval, with fewer presentations in hotter months. Separation in the multivariable model was driven by 44 good-management events, not by treating sample size as the outcome count.

## Conclusion

Good prehospital management of pediatric superficial burns was uncommon in Mazabuka District, with only 13% of children receiving appropriate care. Children brought to health facilities sooner, older children, and those from rural areas were more likely to receive good care, whereas older caregivers and urban residence were associated with poorer care. Community education should emphasise cooling burns under clean, cool running water for 20 minutes, avoiding harmful substances, and promptly seeking medical care. Health education sessions at child health clinics and targeted community outreach for older caregivers and urban households may improve prehospital burn management.

## Data Availability

The data used in this study are available from the corresponding author upon reasonable request.

## Abbreviations

SSA: Sub-Saharan Africa
AOR: Adjusted Odds Ratio
CI: Confidence Interval
IQR: Interquartile Range
LMICs: Low- and Middle-Income Countries

## Acknowledgements

The authors would like to thank the Mazabuka District Health Office and the participating health facilities for their support. We also acknowledge all caregivers who took part in the study and the field teams who made data collection possible. The authors sincerely acknowledge Ms. Emelda Muntali, Malala Mpakani, and Mr. Ronald Fisa for their valuable contributions, support, guidance, and input, which contributed significantly to the completion of this manuscript. Special appreciation is extended to Dr. Nino Paridze for the financial support provided towards this work.

## Author Contributions

DK conceived the study, conducted the statistical analysis, and drafted the manuscript. HER, PKM, CJ, and PM contributed to the study design, interpretation of findings, and critical revision of the manuscript. All authors read and approved the final manuscript.

## Data Availability

The data used in this study are available from the corresponding author upon reasonable request.

## Declarations

### Ethics approval and consent to participate

Ethical approval was obtained from the University of Zambia Biomedical Research Ethics Committee, with additional approval from the National Health Research Authority. Written informed consent was obtained from all participants. UNZABREC REF. No. 6845-2025; NHRA-2425/07/07/2025.

## Consent for publication

not applicable.

